# A conceptual modelling framework for estimating the consequences of psoriatic arthritis disease domains

**DOI:** 10.1101/2021.08.09.21261799

**Authors:** Jacquelyn W Chou, Ervant J Maksabedian Hernandez, David H. Collier, Howard Thom

## Abstract

**Objective:** Characterize psoriatic arthritis (PsA) heterogeneity and develop a conceptual framework to model the clinical and economic value of treatments on key PsA disease domains.

**Methods:** We performed a narrative literature review to identify the key PsA disease domains and outcome measures for these and explore the existing evidence base of clinical and economic evaluations of current PsA treatments on key disease domains. We propose an economic modelling framework that could estimate the economic value of PsA treatments on disease domains.

**Results:** Our literature review identified the following key PsA disease domains: peripheral arthritis, dactylitis, enthesitis, psoriatic spondylitis, skin, and nail lesions, and associated key outcome measures. Our review of the literature, including clinical guidelines, suggests that the evidence base of specific treatment performance on modulating outcomes for a given domain was not strong. We propose an economic modelling framework to estimate the economic value of treatments for addressing specific PsA disease domains. Given existing evidence gaps, we would recommend a cohort-level model with a lifetime horizon, to estimate total social value as well as the cost per outcome and change in cost per symptom avoided. The significant patient heterogeneity in PsA would suggest an individual/patient-level simulation but is likely not feasible with existing data.

**Conclusions:** PsA is a heterogeneous disease, with a number of key disease domains and variable presentation. A better understanding of the performance of treatments on given disease domains would help support patients, providers, payers, HTA’s, and health care policy makers in making personalized treatment decisions.

**Key messages:**

1. Psoriatic arthritis is a heterogeneous disease, with patients presenting a variety of disease manifestations.
2. Better understanding of the interaction between manifestations and treatments could help inform patient and physician decision making.
3. The proposed model framework would help inform value-based, personalized treatment decisions in PsA.

## Background

Psoriatic arthritis (PsA) is an inflammatory joint disease that occurs in many people affected with psoriasis.(1, 2) The mean age at diagnosis has been estimated to be between 40.7 and 52.0 years (median 47.7 years).(2) Patients may experience mobility challenges, lethargy, and limitations on activities of daily living, such as getting in and out of bed, getting in and out of a car, and dressing.(2, 3) PsA is associated with several comorbidities including, but not limited to, diabetes (occurring in 15.9% of psoriatic arthritis patients), hypertension (42.3%), metabolic disease (58.1%), and cardiovascular disease (56.4%).(4, 5)

A meta-analysis carried out by Scotti et al. found a wide variation in the annual prevalence and incidence of PsA; 133 cases for every 100,000 subjects (95% CI, 107–164 every 100,000 subjects) and 83 cases every 100,000 person years (95% CI, 41–167 every 100,000 PY), respectively.(6) Another systematic literature review noted that large differences in the study estimates may be observed among different countries.(7) Additionally, depending on the definitions used (e.g. diagnostic codes, patient self-reporting, rheumatologist diagnosis and classification criteria) prevalence and incidence findings vary substantially.(6)

PsA is a distinctly heterogeneous disease with a number of different symptomatic manifestations. Patients may experience one manifestation alone, or several in combination.(2, 8) Due to the nature of PsA, treatments are tailored to address each patient’s unique symptom profile. It would be beneficial to patients, providers, payers, HTA’s, and health care policy makers to better understand the therapeutic and economic impacts of specific treatments on each disease domain and on various symptom profiles.

Health economic modelling can be utilized to evaluate the cost-effectiveness of treatments in heterogeneous health conditions. With appropriate modelling and data, it is possible to estimate which treatments are the most effective and valuable across a variety of disease manifestations and combinations.(9) Subgroup or individual-level simulations are recommended, by the International Society for Pharmacoeconomics and Outcomes Research (ISPOR) and Society for Medical Decision Making (SMDM), when modelling heterogeneous patient populations.(10)

Previous models in psoriatic arthritis have focused on a general set of outcomes and domains. Broad composite measures of disease activity such as the Psoriasis Area and Severity Index (PASI), which measures body surface area, erythema, induration, and scaling of psoriasis on different areas of the body(11), the American College of Rheumatology joint count (ACR), which includes a 68/66 joint assessment, and Psoriatic Arthritis Response Criteria (PsARC), which also includes a 68/66 joint assessment as well as, the patient’s opinion of their global health and the physician’s global assessment(12) have been used to estimate treatment value in the general PsA patient population.

Prior models, notably the much-cited York model, have not considered the significant heterogeneity of patient populations. Similarly, models conducted for submission to global health technology assessments have lacked inclusion of more granular patient clinical characteristics. For example, in the 2010 National Institute for Health and Care Excellence (NICE) review of etanercept and infliximab for the treatment of PsA, it was noted that none of the submitted models evaluated disease domains beyond psoriasis. The NICE Assessment Group later included joint disease in their evaluation. They found that the utility was driven by the patients’ joint disease response (the Health Assessment Questionnaire (HAQ) response) rather than the skin response (PASI). This result indicates that different disease domains may have variable effects on utility, and thus value.

In the decade since the last PsA review, NICE have not conducted additional reviews to consider new evidence. Similarly, in a recent review of ixekizumab for PsA, the Institute for Quality and Efficiency in Health Care (IQWiG) in Germany focused on the American College of Rheumatology criteria (ACR20) as the primary outcome. Other domain-specific measures of disease activity, including the Leeds Enthesitis Index (LEI), the Leeds Dactylitis Index-Basic (LDI-B), Nail Psoriasis Severity Index (NAPSI), PASI 100, and the Bath Ankylosing Spondylitis Disease Activity Index (BASDAI), were considered as secondary outcomes.(13)

Modern research has recognized the variability of disease presentation among patients with PsA.(11) Aside from the measures considered by IQWiG, there are other instruments that quantify domain-specific disease activity. For example, the Spondyloarthritis Research Consortium Canada (SPARCC) Enthesitis Index hones in on enthesitis disease activity and the Ankylosing Spondylitis Activity Score (ASDAS) captures disease activity involving axial disease.

Current measures of disease domain activity are able to deliver more granular information about patients’ unique symptom manifestations, adding further support for pervasive disease heterogeneity. Economic model frameworks must be adapted to accommodate the most robust, clinically relevant data available.

In this paper, we propose an economic modelling framework to estimate the value of individual treatments for managing simple and complex presentations of PsA, incorporating one or more disease domains. In order to do this, we first look into the current data availability in disease outcomes for PsA. This model aims to more accurately capture and reflect real world variations in the experience of PsA treatments.

## Methods

We conducted a narrative literature review using PubMed and Tufts Cost-Effectiveness Analysis Registry (CEA Registry) to answer the following key questions:

1. What are the key disease domains that psoriatic arthritis patients manifest?
2. What is the prevalence of each domain?
3. What is the overlap between domains?
4. How does the value of treatment differ across disease domain subgroups with psoriatic arthritis?
5. Do psoriatic arthritis treatments have a more significant impact on one disease domain subgroup than on others?
6. How do health-related impacts translate to value among the psoriatic arthritis subgroups?

To identify information regarding key disease domains for questions 1 through 3, we searched PubMed using combinations of the following search terms: psoriatic arthritis, subtypes, domains, manifestations, epidemiology, and natural history. To identify treatment and outcomes related information as well as measures of the key disease domains captured in clinical trials for questions 4 and 5, we searched for PubMed using the following list of biologic treatments for PsA to ensure capture of the clinical benefit of the more recent innovations for psoriatic arthritis : abatacept (Orencia), adalimumab (Humira), certolizumab (Cimzia), etanercept (Enbrel), golimumab (Simponi), infliximab (Remicade), ixekizumab (Taltz), secukinumab (Cosentyx), tofacitinib (Xeljanz), apremilast (Otezla), ustekinumab (Stelara) looking at studies published in 2000 or later. Lastly, to identify existing cost-effectiveness models evaluating the PsA disease domains to answer question 6, we searched the Tufts CEA Registry for “psoriatic arthritis.”

We reviewed English language articles related to key PsA disease domains, common instruments used to assess patients with PsA disease domains, economic models on PsA, and conceptual framework articles for multi-state Markov models in PsA or similar disease areas to identify common modeling approaches for framework development. Model frameworks, clinical trials, observational studies, and treatment guidelines were included in this review. Iterative search strings were used, with titles screened up until the 100th result, or until remaining titles were not relevant to the present study. The abstracts of potentially relevant titles were then read, after which the full text of the article was assessed for inclusion. After identifying relevant papers, we also conducted citation tracing to ensure key foundational papers were identified.

We used the identified literature to develop the model framework in three primary ways. First, papers identified to answer questions 1 through 3 helped us determine the key components for a model in PsA looking to capture heterogeneous disease domains. Second, the papers identified to answer questions 4 through 6 helped to identify potential approaches for a PsA model. Lastly, we compared the findings for questions 1 through 3 with those from questions 4 through 6 to determine what gaps and limitations current models had and whether models in other disease areas could offer suggestions for how to fill those gaps.

Because the study collected only published and publicly available secondary data and did not involve the collection, use, or transmittal of individually identifiable data, institutional review board approval to conduct this study was not necessary.

## Results

### Literature conceptualizing psoriatic arthritis

As a result of our search and citation tracing, we identified three key sources for defining relevant PsA domains: American College of Rheumatology & National Psoriasis Foundation (ACR/NPF) guidelines(14), Group for Research and Assessment of Psoriasis and Psoriatic Arthritis (GRAPPA) treatment guidelines(15), and Moll and Wright (1973)(16).

In their seminal paper, Moll and Wright divided psoriatic arthritis into ‘psoriasis symptoms’ and ‘rheumatoid arthritis symptoms’.(16) Since then, psoriatic arthritis has been recognized as a unified disease that presents in several manifestations. The ACR/NPF guidelines identify four disease manifestations: peripheral arthritis, dactylitis, enthesitis, and psoriatic spondylitis. Nail lesions and skin involvement, specifically red and crusty skin patches, are included in the definition of active PsA. The GRAPPA guidelines recommend that assessment of patients requires consideration of all major disease domains including peripheral arthritis, axial disease, enthesitis, dactylitis, psoriasis, and nail disease.(15)

The Corrona Registry, a Psoriatic Arthritis data warehouse containing rich clinical detail, appeared several times in our targeted literature review. The Corrona PsA/SpA registry has collected in-depth information on patients with PsA since 2001 including clinical characteristics, clinician reported outcomes, and patient reported outcomes.(17)

Corrona registry publications reported that among the PsA patients, 65% had more than one disease domain manifestation, 24% had one disease domain manifestation, and 11% had no domains manifestation.(18) Of those patients with at least one PsA disease domain manifestation in the Corrona registry, 1814 (78.4%) presented with skin disease, 1523 (65.8%) with peripheral arthritis, 1042 (45.0%) with nail psoriasis, 539 (23.3%) with enthesitis, 319 (13.8%) with axial disease, and 235 (10.2%) with dactylitis.(18)

Psoriatic arthritis disease presentation varies from patient to patient, making treatment decisions and outcomes variable.(8) The most prevalent domain is skin involvement, also called psoriasis – a condition that causes dry, erythematous, raised skin patches with slivery scale, and chronic itching.(18, 19) Peripheral arthritis is also common, and causes swelling, pain, and/or stiffness in the hands, feet, knees, or areas other than the spine. Pain and/or stiffness of the joints in the spine and/or buttocks region is categorized as axial disease or spondylitis. Enthesitis is swelling, pain, and/or stiffness where tendons and ligaments connect to the bone. Dactylitis describes diffuse swelling of the fingers and/or toes due to a combination of tendonitis, arthritis, and enthesitis. Lastly, nail psoriasis can be one or more findings of pin-sized indentations, nail thickening, ridges, and onycholysis of the nail.

We found that disease activity measurements have become reflective of the heterogeneity in the PsA patient population. In addition to general measures of inflammation and composite measures, domain-specific measures are widely available. Peripheral arthritis is measured by the American College of Rheumatology 20% response criteria (ACR20).(11) Clinical measures of axial disease include the Bath Ankylosing Spondylitis (AS) Disease Activity Index (BASDAI) and the AS Disease Activity Score (ASDAS).(11) The Spondyloarthritis Research Consortium Canada (SPARCC) Enthesitis Index and the Leeds Enthesitis Index have been used to measure enthesitis disease activity.(11) The Leeds Dactylitis Index (LDI) measures dactylitis disease activity.(11) Nail psoriasis is usually gauged with a visual inspection of the nailbeds while, for studies, the Nail Psoriasis Severity Index (NAPSI) or modified NAPSI (m-NAPSI) is used to measure nail involvement.(11) Given our emphasis on discrete domain specificity, we elected not to include composite scores that combine measures across domains in our review.

Current ACR/NPF treatment guidelines and recommendations involve selecting the best treatment for the patient’s symptoms, severity, and comorbidities as assessed by a physician.(14) These considerations are critical to effective therapeutic management of disease, and are common among several treatment guidelines found during this literature review.(14, 20, 21) The ACR/NPF have developed a detailed guideline for the treatment of PsA. The guidelines divide treatments into three buckets: (1) Non-pharmacologic, (2) symptomatic, and (3) pharmacologic. Non-pharmacologic treatments for PsA include physical therapy, occupational therapy, smoking cessation, weight loss, massage therapy and exercise. Symptomatic treatments identified in the guidelines are Non-steroidal anti-inflammatory drugs (NSAIDs) and glucocorticoids. ^21^ Largely, the guidelines focus on recommendations among the therapies in the pharmacologic treatment bucket, outlined in **Table 1**.

**Table 1.** Identified outcomes in PsA disease and the GRAPPA treatment guidelines specifying which disease domains are treated using specific treatments.

| Treatment | Target | Peripheral Arthritis | Skin disease | Nail disease | Psoriatic Spondylitis | Dactylitis | Enthesitis |
| --- | --- | --- | --- | --- | --- | --- | --- |
| secukinumab | Cytokines: IL-17A | ✓ | ✓ | ✓ | ✓ | ✓ | ✓ |
| adalimumab | TNF | ✓ | ✓ | ✓ | ✓ | ✓ | ✓ |
| etanercept | TNF | ✓ | ✓ | ✓ | ✓ | ✓ | ✓ |
| apremilast | small-molecule inhibitor that modulates inflammatory cytokine production via inhibition of phosphodiesterase-4 (PDE-4 inhibitor) | ✓ | ✓ | ✓ |  | ✓ | ✓ |
| certolizumab | TNF | ✓ | ✓ | ✓ | ✓ | ✓ | ✓ |
| golimumab | TNF | ✓ | ✓ | ✓ | ✓ | ✓ | ✓ |
| infliximab | TNF | ✓ | ✓ | ✓ | ✓ | ✓ | ✓ |
| ustekinumab | Cytokines: IL-12 and IL-23 | ✓ | ✓ | ✓ |  | ✓ | ✓ |

**Table 2.**
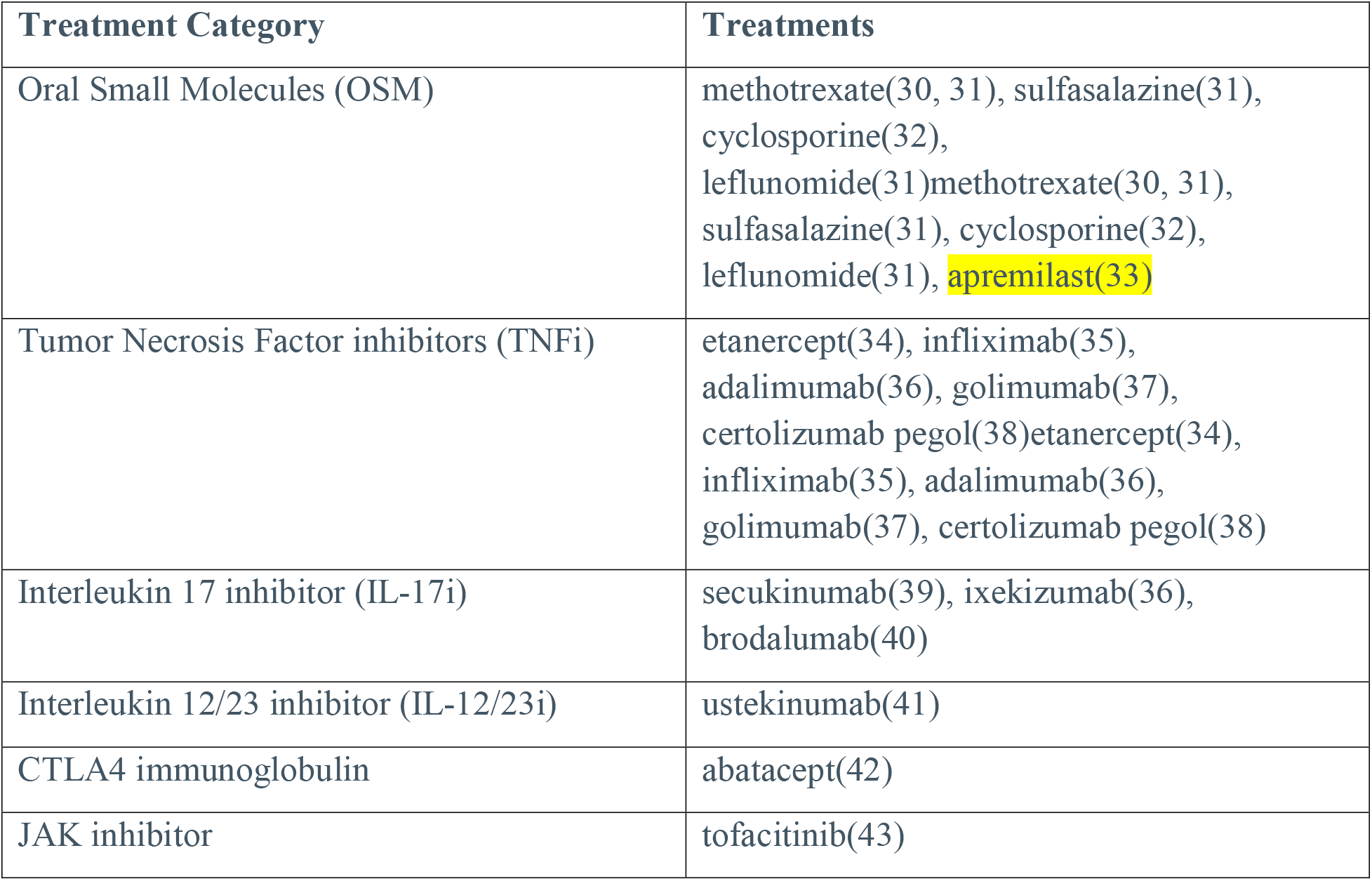
Pharmacologic treatments included in ACR/NPF’s PsA therapy review and guidelines.

| Treatment Category | Treatments |
| --- | --- |
| Oral Small Molecules (OSM) | methotrexate(30, 31), sulfasalazine(31), cyclosporine(32), leflunomide(31)methotrexate(30, 31), sulfasalazine(31), cyclosporine(32), leflunomide(31), **apremilast(33)** |
| Tumor Necrosis Factor inhibitors (TNFi) | etanercept(34), infliximab(35), adalimumab(36), golimumab(37), certolizumab pegol(38)etanercept(34), infliximab(35), adalimumab(36), golimumab(37), certolizumab pegol(38) |
| Interleukin 17 inhibitor (IL-17i) | secukinumab(39), ixekizumab(36), brodalumab(40) |
| Interleukin 12/23 inhibitor (IL-12/23i) | ustekinumab(41) |
| CTLA4 immunoglobulin | abatacept(42) |
| JAK inhibitor | tofacitinib(43) |

Available evidence is inconclusive regarding the efficacy of oral small molecules (OSMs) in management of PsA, whereas there is moderate-quality evidence of the benefits of Tumor Necrosis Factor inhibitors (TNFi) biologics. ^21^ In particular, TNFis have a marked impact on the prevention of disease progression and joint damage. In general, biologic monotherapy is preferred to combination therapy with methotrexate.^21^

The ACR/NPF guidelines explicitly state that significant challenges exist in evaluating and recommending treatments in a heterogeneous population with a wide range of possible symptom profiles. There is agreement that the current treatment paradigm is limited in providing more tailored treatment decisions. While availability of domain specific evidence has accumulated,(18) there is no single algorithm that is able to capture heterogeneity in the presentation and course of PsA.

### Literature on comparative effectiveness

In our literature search of the Tufts CEAR database, we identified 14 papers on the topic of PsA. Only 3 of those papers made mention of a range of psoriatic arthritis disease symptoms in their economic model. Full text review of those 3 papers only identified one, Olivieri (2008) (22), that discussed the relevance of evaluating treatment outcomes for specific disease domains. However, the Olivieri (2008) article did not explicitly include a comparative evaluation of psoriatic treatments on particular disease domains, limiting its utility as a model for examining treatment effects on heterogeneous PsA disease domains.

We did not identify any other economic models that provided a comparative evaluation of treatment performance on PsA disease domains. A more granular understanding of the interaction between disease domains and treatments would benefit both providers and patients in choosing the appropriate treatment. However, we did not identify a clinical trial or network meta-analysis providing a comprehensive comparison of PsA treatments with specific disease domain outcomes, suggesting that such granularity is not available in PsA data. Incorporating disease subgroups and treatment interactions into an economic model would provide support for more personalized, value-based treatment decisions. Unfortunately, the lack of high-quality data prevents us from creating a fully functional model that captures the heterogeneity of this disease and the comparative effectiveness of treatments. Therefore, we propose a modelling framework in preparation for a future when these data are available.

### Proposed modelling framework

We propose a cohort-level economic model with a lifetime horizon, given the chronic nature of PsA. The components of our conceptual model for PsA outcomes are illustrated in

Figure 1. The influence of these components on treatment, disease outcomes, and health economic outcomes is illustrated in Figure 2. The dependent variables in

**Figure 1.**
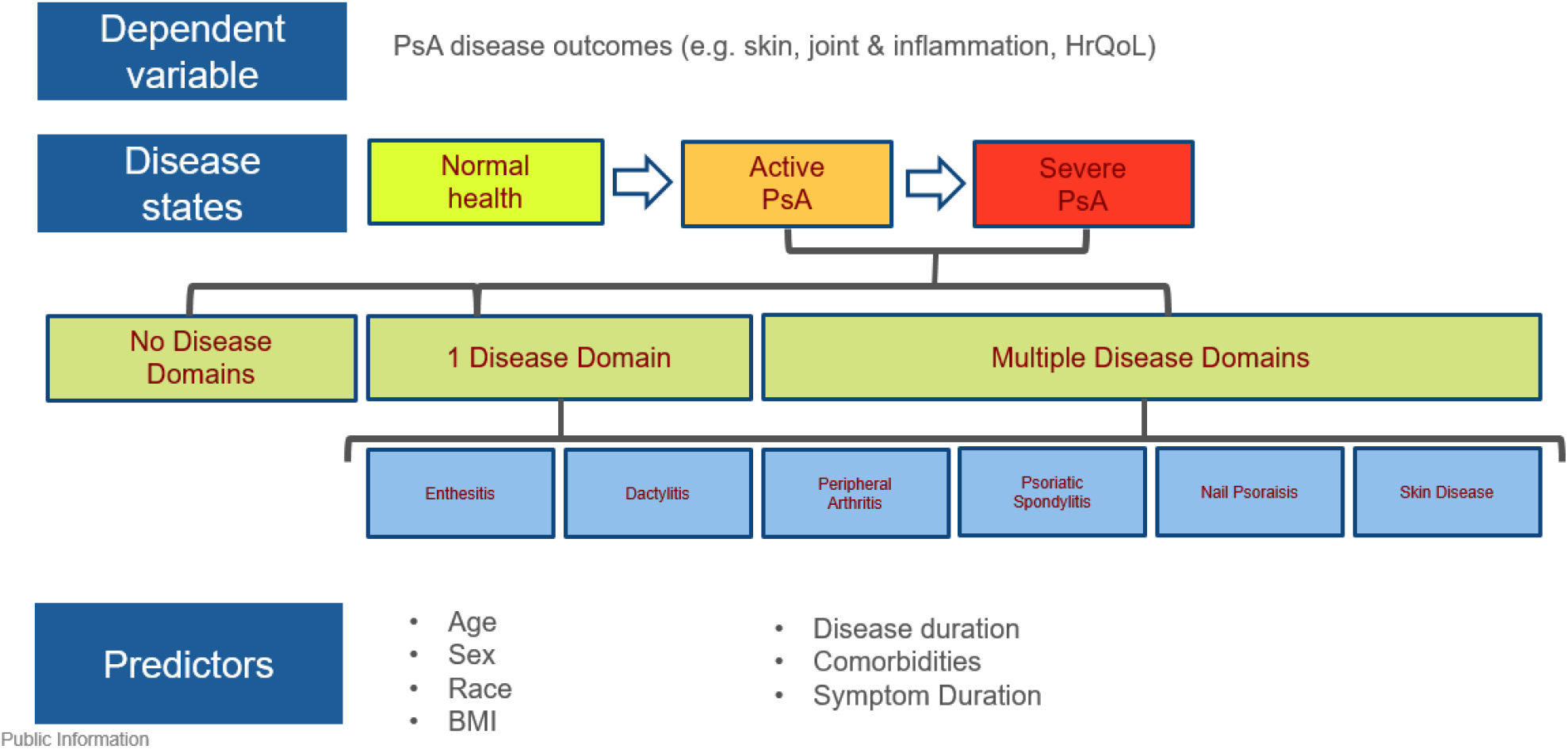
Components of conceptual model for PsA outcomes

**Figure 2.**
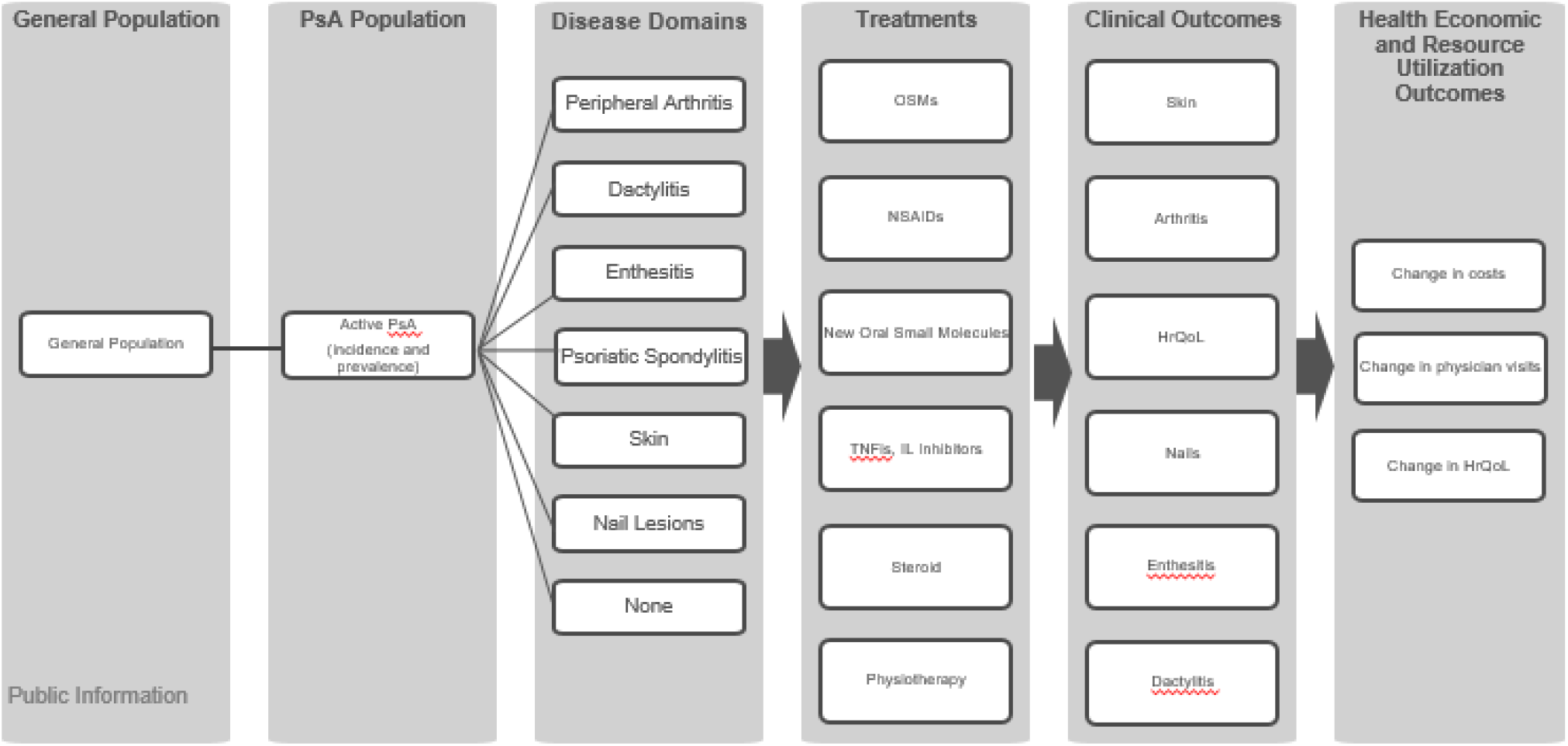
Conceptual model for influence of PsA disease on treatment, clinical outcomes, and health economic outcomes. PsA - psoriatic arthritis; OSMs – oral small molecules; NSAIDs – non-steroidal anti-inflammatory drugs; TNFis – tumor necrosis factor inhibitors; IL – interleukin; HRQoL – health related quality of life;

Figure 1 are PsA disease outcomes (i.e. those related to skin or join inflammation, enthesitis, etc.) which will determine costs and quality of life. These outcomes depend on health states and individual predictors (age, sex, race, body mass index (BMI), disease duration, comorbidities, and symptom duration). At the highest level, we categorize the general population into normal health (i.e. no PsA) and active PsA. Aligning with the overlap of our three key sources on disease manifestations (ACR/NPF, GRAPPA, Moll and Wright), we divided active PsA into six key disease domains: peripheral arthritis, dactylitis, enthesitis, psoriatic spondylitis, skin involvement, and nail lesions. PsA patients may have none, one, or many of these disease domains, as indicated by the Corrona registry.(18)

As recommended by treatment guidelines, and reported in practice, treatment decisions are made on a patient-by-patient basis in response to the active disease domains and severity. This is indicated by the influence of disease domains on treatment selection, as depicted in Figure 2. We have categorized treatments into OSMs, NSAIDs, new OSMs, TNFis/Interleukin inhibitors (IL), steroids, and physiotherapy. These non-overlapping therapies are recommended for the treatment of PsA in the ACR/NPF guidelines. (18)

PsA severity, disease domains, treatments, and individual predictors influence the PsA specific clinical outcomes relating to skin, arthritis, nails, enthesitis, dactylitis, and health related quality of life (HRQoL). As described previously, there are a wide range of measures possible for each of these outcomes. Peripheral arthritis is measured by ACR20; axial disease by BASDAI and/or ASDAS; enthesitis by the SPARCC Enthesitis Index and/or the Leeds Enthesitis Index; dactylitis by the Leeds Dactylitis Index; nail psoriasis is usually gauged with a visual inspection of the nailbeds or by using the NAPSI or m-NAPSI. Meanwhile, HRQoL can be measured by the short form 36 (SF-36), the health assessment questionnaire disability index (HAQ-DI), functional assessment of chronic illness therapy fatigue (FACIT-F)(23) and Dermatology Life Quality Index (DLQI), as well as many others. Our framework is neutral to the measure used in practice.(11)

The final influence step in our conceptual framework is from PsA clinical outcomes to health economic and resource utilization outcomes. As indicated by our review of the literature, the primary outcomes in this step are changes in HRQoL, physician visits, and costs. HRQoL refers to all aspects of quality of life and may be wider than indicated by a PsA-specific HRQoL measure; for example, though psychological distress is included, mental health is not included in the HAQ-DI. We include physician visits, as PsA patients are usually seen in outpatient care, as opposed to inpatient care.(24) Additionally, frequency of physician visits functions as a proxy for disease activity. Costs include both health (e.g. cost of physician visit, medication, physiotherapy, occupational therapy) and non-health (e.g. reduced quantity of employment, greater vehicular transport use to overcome mobility issues) costs.

Figure 3 demonstrates one example of flow through in the proposed model. In this case, the PsA population selected in the model has a domain presentation of dactylitis. Three treatments (OSMs, TNFis, and IL inhibitors) have been identified for comparison in the treatment of dactylitis. The anticipated outcomes of the model would show differences in social value calculated as quality-adjusted life years, costs per grouped outcome, and change in cost per symptom avoided across treatments for the selected subgroup of PsA patients with dactylitis.

**Figure 3.**
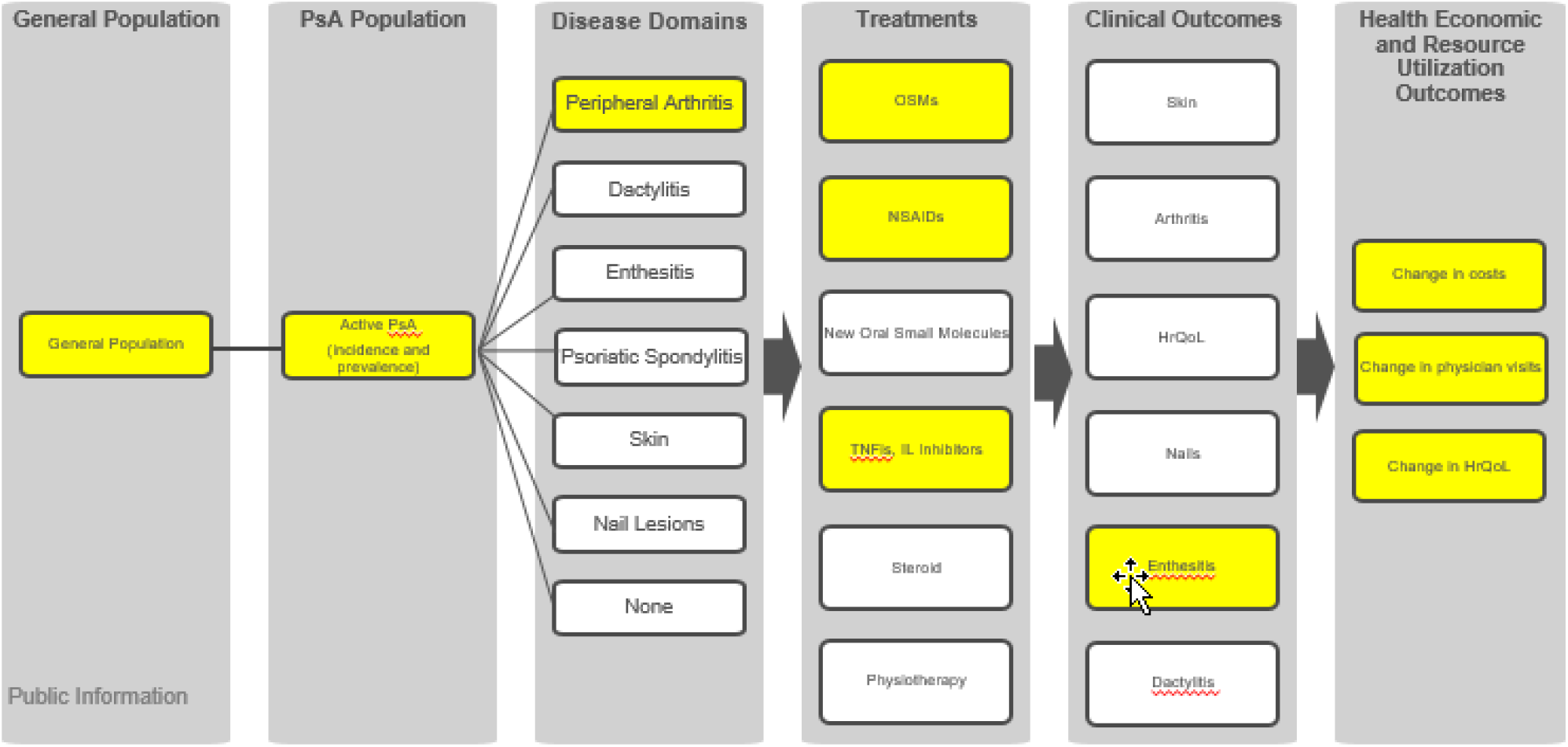
Example of flow-through our conceptual model PsA - psoriatic arthritis; OSMs – oral small molecules; NSAIDs – non-steroidal anti-inflammatory drugs; TNFis – tumor necrosis factor inhibitors; IL – interleukin; HRQoL – health related quality of life;

### Data sources and implementation

The proposed modelling framework will be able to provide more granular health economic outcomes than previous models. However, implementation would require richly detailed evidence sources and careful model programming.

Current and granular data, offered by sources such as the Corrona registry(18) or a network meta-analysis, on domain- and treatment-specific inputs would be needed to populate the model (Figure 1). Patient-level data on the prognosis predictors (Figure 1) age, sex, race, body mass index (BMI), disease duration, comorbidities, and symptom duration would also be required. Treatment effects are unlikely to be completely captured by registry data and there are no head-to-head trials of all treatment groups on all PsA outcomes, so indirect treatment comparisons would be needed. Furthermore, treatment effects differ by domain and the domains are correlated, so a multiple outcomes indirect treatment comparison would be instituted.(25) This comparison could be accomplished by Bayesian network meta-analysis. However, patient populations and other aspects of clinical trials tend to vary considerably. In practice, Bayesian meta-analysis may not help researchers populate a model that tried to capture the heterogeneity in PsA.(26)

Further input data would include healthcare resource utilization and costs to the healthcare system. Demographics and clinical characteristics should be sourced from relevant clinical trials and from real world studies, where possible.

Our modelling framework lends itself to a state-transition modelling implementation. We would suggest a multistate Markov model structure, in which one or more disease domains can be evaluated. State occupancy would be used over time to estimate health economic and resource utilization outcomes. An individual patient simulation would best model heterogeneous disease domains, treatment responses, and individual risk factors.(27) However, this is contingent on data availability and a cohort model, or series of cohort models on different subgroups of patients defined by disease domains, may be more feasible. A cost-utility analysis would be conducted but the granularity of our model would also enable cost-consequence analysis, which does not require a judgement of the relative importance of the final outcomes indicated in Figure 2.

## Discussion

Psoriatic arthritis is a heterogeneous disease, with patients presenting a variety of disease manifestations and combinations. Therapeutic efficacy can vary widely, interacting with patients’ unique symptom profiles to produce varied estimates of value. Past models have not generally reflected this complexity, more commonly using composite measures of disease activity rather than modeling the impacts on individual disease domains. This may be in part due to the lack of comparative evidence of treatment impacts on individual disease domains. While each of the disease domains has an existing outcome to measure activity for the domain, these measures were not uniformly employed across all clinical trials for all treatments.

Similarly, current treatment guidelines largely focus on general disease and treatment history, rather than specific symptom profiles and disease domain responses. Many measures of disease response are driven by reductions in more common disease domains, such as psoriasis and peripheral arthritis. This may again be due to the lack of evidence on specific outcomes in clinical trials for available PsA treatments.

In the PsA review, the NICE Committee noted the importance of registries in collecting data and supported including outcomes specific to PsA so that specific information about treatments for psoriatic arthritis can be captured.(28) New data systems, like the Corrona Registry, have adapted to collect more granular data from patients with PsA. This type of registry data provides a better representation of disease heterogeneity. Additionally, these registries have the potential to provide information on the response patients have on various disease domains depending on the treatment they are using. This would open up new opportunities for analysis and evaluation of treatment responses. A more granular understanding of the interaction between disease domains and treatments would benefit both providers and patients.

Alternatively, high-quality network meta-analyses and indirect treatment comparisons could leverage existing published clinical trials to develop comparative effectiveness data of treatments on the varying disease domains. However, the inconsistency with which clinical trials measured the various disease domains and the exact measure used for a given disease domain, along with the heterogeneity present in NMAs, might result in incomplete networks on the key domains of interest. (29)Greater alignment in the PsA disease area regarding which disease domains are central as well as the standard set of measures for each disease domain would result in a more uniform set of outcomes across clinical trials, enabling more informed comparative analyses of effectiveness. There is promise for the future as the Group for Research and Assessment of Psoriasis and Psoriatic Arthritis (GRAPPA) and Outcome Measures in Rheumatology (OMERACT) are collaborating to develop a Core Outcome Measurement Set for Psoriatic Arthritis (PsA).(29) It remains to be seen whether their recommendations will be adopted by clinical trials so that harmonization can be accomplished.

In order to actualize the proposed model, there are several steps that must be executed. The authors would recommend further real-world data analyses combined with a systematic literature review and network meta-analysis to better establish natural disease progression and correlation with outcomes. Furthermore, the authors would recommend a study to demonstrate the comparative effectiveness of treatments on each domain or combination of domains.

Given the progress in data collection, there are now building blocks to bridge the gap between disease response and treatment options. Incorporating disease subgroups and treatment interactions into an economic model would provide support for more personalized, value-based treatment decisions. The model framework proposed in this paper would offer the flexibility to examine disease domain profiles and treatment value simultaneously in the PsA population once the data becomes available.

## Data Availability

Qualified researchers may request data from Amgen clinical studies. Complete details are available at the following: http://www.amgen.com/datasharing.

